# Mendelian randomisation analyses of eosinophils and other blood cell types in relation to lung function and disease

**DOI:** 10.1101/2020.07.09.20148726

**Authors:** Anna L Guyatt, Catherine John, Alexander T Williams, Nick Shrine, Nicola Reeve, SpiroMeta consortium, Ian P Hall, Louise V Wain, Nuala A Sheehan, Frank Dudbridge, Martin D Tobin

## Abstract

**Background:** Eosinophils are granulocytes associated with airway inflammation in respiratory disease. Eosinophil production and survival is controlled by interleukin-5: anti-interleukin-5 agents reduce asthma and COPD exacerbation frequency, and response correlates with baseline eosinophil counts. However, causal relationships between eosinophils and other respiratory phenotypes are less studied.

**Methods:** We investigated causality between eosinophils and: lung function, acute exacerbations of COPD (AECOPD), asthma-COPD overlap (ACO), moderate-to-severe asthma, and respiratory infections. We performed Mendelian randomization (MR) using 151 genetic variants from genome-wide association studies of blood eosinophil counts in UK Biobank/INTERVAL, and respiratory data from UK Biobank, using MR methods relying on different assumptions for validity. Multivariable MR using eight blood cell type exposures was performed for outcomes showing evidence of causation by eosinophils.

**Findings:** There was evidence that higher eosinophils reduce FEV_1_/FVC and FEV_1_ (weighted median estimator, SD change FEV_1_/FVC per SD eosinophils: −0.054 [95%CI −0.078,−0.029]. There was also evidence that eosinophils cause ACO (weighted median OR 1.44 [95%CI 1.19,1.74]), and asthma (weighted median OR 1.50 [95%CI 1.23,1.83]). Multivariable MR for FEV_1_/FVC, FEV_1_, ACO and asthma suggested that eosinophils were the cell type with the most important effect. Causal estimates of individual variants were heterogeneous, which may arise from pleiotropy.

**Interpretation:** We found evidence that eosinophils reduce lung function, and increase ACO and asthma risk, on average over the set of genetic variants studied. Eosinophils appear to be causal determinants of fixed airflow obstruction among individuals with features of both asthma and COPD.

**Funding:** Wellcome, BHF, MRC, BBSRC CASE studentship with GSK, GSK/BLF.

## Research in Context

### Evidence before this study

High eosinophils are observed in some individuals with asthma and COPD. Anti-IL5 and anti-IL5R drugs, such as mepolizumab and benralizumab, respectively, are currently licensed for treatment of severe eosinophilic asthma, and response correlates with baseline eosinophil levels. Clinical trials of anti-IL5 agents in COPD have reported reductions in blood and sputum eosinophil counts, but not reproducible reductions in COPD exacerbations, and any clinical improvement in COPD has been hypothesised to be smaller than that in asthma, and similarly related to the degree of eosinophilic inflammation.

### Added value of this study

We performed Mendelian randomization (MR) analyses, using genetic variants that predict blood cell counts, to investigate causation between eosinophils and several quantitative lung function traits and clinical respiratory outcomes, encompassing both fixed and reversible airflow obstruction. Where there was evidence of causation by eosinophils, we explored whether other blood cell types may have contributed to this association, since blood cells counts are correlated. Overall, our aim was to provide a comprehensive assessment of the causal role of blood eosinophil counts in respiratory health and disease.

### Implications of all available evidence

Although clinical trials of anti-IL5 agents have not directly tested the causal role of eosinophils in asthma and COPD, they are hypothesised to act via a reduction in eosinophil counts. We found evidence for causality of eosinophils on FEV_1_ and FEV_1_/FVC (used in the diagnosis of COPD), on asthma-COPD overlap, and on asthma. We did not find evidence for eosinophils causing COPD exacerbations, but cannot rule out a small effect. Whilst the average effect of lowering eosinophils is to improve certain respiratory phenotypes we noted heterogeneity in our causal estimates from individual genetic variants, suggesting pleiotropic effects of SNPs affecting eosinophil levels. Taken together with prior evidence, our findings suggest that clinical trials of eosinophil lowering agents are warranted in patients with history of both asthma and COPD.

## Introduction

Eosinophils are proinflammatory granulocytes associated with symptom severity and exacerbation frequency in patients with asthma and chronic obstructive pulmonary disease (COPD).^2-4^ The degree of eosinophilia in these obstructive lung diseases is variable: whilst eosinophil inflammation brought about by allergic sensitisation has been considered characteristic of asthma, not all patients with asthma have eosinophilia.^2,5^ Moreover, whilst airway inflammation in COPD is typically mediated by neutrophils, a subset of individuals with COPD have raised eosinophil counts.^2,6^

The production and survival of eosinophils is regulated by interleukin-5 (IL-5), and anti-IL5 therapies (e.g. mepolizumab, and the anti-IL5RA agent, benralizumab) are now licensed in many countries for the treatment of severe eosinophilic asthma.^7-13^ The decision to treat asthma with these drugs is currently based upon blood eosinophil count,^2^ since post-hoc analyses of clinical trials stratified by eosinophil levels have shown increased efficacy of mepolizumab for treating severe asthma in those with higher baseline eosinophils.^3^ Results from Mendelian randomization (MR) analyses have also provided evidence for a causal role of eosinophils in asthma.^14^ MR analyses use genetic variants as instrumental variables (IVs) to investigate causality between exposure and outcome, and under certain assumptions may obviate some of the problems of traditional observational epidemiology (reverse causation and confounding), permitting causal inference.

In addition to asthma, blood eosinophil counts are associated with quantitative measures of lung function in healthy populations (i.e. including individuals without asthma).^15^ However, causality of these associations has yet to be established, with the only previous MR of lung function being of small sample size, with imprecise effect estimates precluding confident inference.^16^ Moreover, causal effects of eosinophils on other respiratory phenotypes, such as asthma-COPD overlap (ACO), and respiratory infections are yet to be investigated. Diagnosis of COPD is made by spirometry if the ratio of the forced expiratory volume in one second (FEV_1_) to the forced vital capacity (FVC), FEV_1_/FVC, is <0.7, with airflow obstruction being graded according to predicted values of FEV_1_. Therefore, studying the role of eosinophils in determining quantitative lung function (i.e. lung function measured as continuous traits) is a powerful way of understanding their role in the development of fixed airflow obstruction such as is observed in COPD.^17,18^ Understanding the causal relationship between eosinophils and fixed airflow obstruction is particularly pertinent given recent interest in the potential use of mepolizumab in COPD.^10-13^

We undertook two-sample MR analyses using summary-level genome-wide association study (GWAS) data to assess causality between eosinophils and respiratory traits and conditions encompassing fixed and reversible airflow obstruction, using genetic variants associated with blood eosinophil counts in the largest GWAS to date as IVs.^14^ First, we investigated a causal effect of eosinophils on three quantitative lung function traits measured by spirometry, and four clinical respiratory phenotypes (moderate-to-severe asthma, acute exacerbations of COPD [AECOPD], ACO, and respiratory infections). We used three MR approaches that rely on different assumptions for validity, and followed-up traits showing evidence of causality to assess whether the instrumental variables affected lung function purely via eosinophil counts and not via other blood cell types. Overall, our aim was to provide a comprehensive assessment of the causal role of blood eosinophil counts in relation to respiratory health and disease.

## Methods

We assessed causal associations between eosinophils and other blood cell counts in relation to multiple respiratory outcomes, using MR.^1,19^ Briefly, MR involves using genetic variants (here single-nucleotide polymorphisms, SNPs), as instrumental variables (IVs) for an exposure of interest, in this case eosinophil counts. This is done by comparing the magnitude of the effect of the SNPs on the outcome to the effect of the SNPs on the exposure.^1,19^ All analyses reported here are two-sample MR analyses, since SNP-exposure and SNP-outcome associations were extracted from different (yet overlapping^20^) samples. The core assumptions of MR for inferring causality between an exposure and outcome are described in **Box 1**. Additional assumptions required for accurate point estimation are discussed in the **Extended Methods**, and elsewhere.^21^

### Box 1 Mendelian randomization: core assumptions

#### Assumptions

Mendelian randomization may be used to test for causality between an exposure (e.g. eosinophils) and outcome (e.g. FEV_1_/FVC), if the following core assumptions hold:

i. the genetic variation (single nucleotide polymorphisms in this work) used as instrumental variables (IVs) are associated with the exposure of interest;
ii. the genetic variants are not associated with unobserved confounders of the exposure-outcome association (short dashed arrow). Genetic variants are allocated randomly at conception (Mendel’s law of independent assortment) and so typically should not be associated with these confounding variables;
iii. association between the genetic variants and the outcome is via the exposure, and not via an alternate pathway (i.e. there is no ‘horizontal pleiotropy’, see long dashed arrow). Whilst difficult to verify, reassurance that this assumption holds can be provided using biological knowledge of how the SNP functions, and by checking whether multiple MR methods, each relying on different assumptions for validity, give consistent results (known as triangulation).^1^

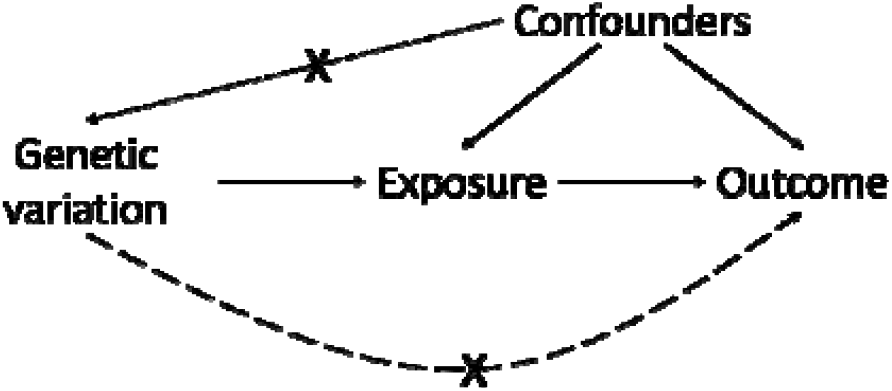

All GWAS datasets analysed included participants from UK Biobank,^22^ incorporating other studies where available. All GWAS included were of individuals of European ancestry. Datasets are summarised here, and descriptions of covariate adjustments, and exposure-outcome GWAS overlap are given in the **Extended Methods**.

### Exposure GWAS data sets (blood cell parameters)

We used summary-level data from a previously published GWAS of blood cell counts ^14^ in the initial release of UK Biobank genetic data (around one third of all participants), plus the INTERVAL study (eosinophil GWAS N=172,275, see **Extended Methods**).^14^ These included counts of eosinophils, basophils, neutrophils, monocytes, lymphocytes, platelets, red blood cells and reticulocytes.

### Outcome GWAS data sets (respiratory outcomes)

#### Quantitative lung function GWASs

We used published summary-level data from three GWAS of quantitative lung function (FEV_1_, FVC and FEV_1_ /FVC), undertaken in 400,102 individuals.^18^

#### Clinical disease GWAS

##### Moderate-to-severe asthma

We used a published GWAS of moderate-to-severe asthma comprising 5,135 cases and 25,675 controls, sampled from the initial release of UK Biobank, supplemented with external cases.^23^

##### Acute exacerbations of COPD

We defined acute exacerbations of COPD (AECOPD) in UK Biobank; the eligible sample was restricted to individuals withFEV_1_/FVC<0.7. From this subset, exacerbation cases (N=2,771) had an ICD-10 code for AECOPD or a lower respiratory trait infection in the hospital episode statistics data provided by UK Biobank (**Supplementary Table 1**. Controls (N=42,052) were people with COPD without a recorded ICD-10 AECOPD code. Associations were adjusted for age (at recruitment), age^2^, sex, smoking status (ever/never), genotyping array and 10 ancestry principal components.

##### Asthma-COPD overlap (ACO)

We defined ACO cases in UK Biobank (N=8,068) as individuals with a self-report of a doctor diagnosis of asthma, and both FEV_1_/FVC<0.7 and FEV_1_<80% predicted at any study visit. Controls (N=40,360) were selected in a ratio of approximately five controls to one case from participants reporting no asthma or COPD, and with FEV_1_ >80% predicted and FEV_1_/FVC>0.7. Associations were adjusted for age (at recruitment), sex, smoking status, and 10 ancestry principal components.

##### Respiratory infections

We defined respiratory tract infections requiring hospital admission in UK Biobank, using a wide range of ICD-10 codes (**Supplementary Table 2**). Cases were individuals with one or more admission for respiratory infections (N=19,459). Controls had no admissions for respiratory infections and were selected in a ratio of approximately five controls to one case (n=101,438 after exclusions). Associations were adjusted for age (at recruitment), age^2^, sex, smoking status, genotyping array, and 10 ancestry principal components.

### Statistical methods

#### Univariable MR analyses of eosinophils and all respiratory traits and diseases

We performed seven separate MR analyses of eosinophils on all outcomes, including three quantitative lung function traits (FEV_1_, FVC, and FEV_1_/FVC); and four clinical disease phenotypes (asthma, AECOPD, ACO and respiratory infections) utilising genetic IVs selected from_18_. The selection of genetic IVs and the harmonisation of SNP-exposure and SNP-outcome datasets is detailed in the **Extended Methods**. A total of 151 eosinophil IVs were used for this analysis. The primary analysis used the inverse-variance weighted (IVW) MR method, and we checked sensitivity to condition (iii) described in Box 1 using sensitivity analyses, conducted using MR-Egger regression,^24^ and the weighted median estimator^25^ (see **Extended Methods** for details, including the different assumptions that the methods rely upon for validity). Further sensitivity analyses: i) restricted to non-UKB FEV_1_/FVC GWAS data, to assess sensitivity to sample overlap, and ii) restricted to FEV_1_/FVC GWAS data in UKB, excluding individuals with asthma.

#### Multivariable MR analyses of multiple blood cell types and respiratory outcomes

Since SNPs affecting eosinophils also affect other blood count types,^14^ we used multivariable MR in order to estimate the influence of multiple cell types on respiratory outcomes, after conditioning on the effects of the SNPs on other cell types. Multivariable MR analyses were run for respiratory outcomes that showed evidence of eosinophil causation in the IVW MR analyses above, and that had broadly consistent effect estimates in the weighted median and MR-Egger analyses. We also performed an analysis of FEV_1_/FVC in UKB (excluding asthma cases).

There were 1166 SNPs associated with at least one of eight blood count traits reported by Astle *et al*^14^ at a genome-wide threshold. SNPs were LD clumped, and effect sizes were extracted from each blood cell trait GWAS, and from the outcome GWASs. Effects for 318 clumped SNPs were successfully harmonised, i.e. so effect sizes for the SNP-exposure and SNP-outcome effects corresponded to the same allele (**Supplementary Table 3, Extended Methods**).

To implement multivariable MR, we used the mv_multiple() function of the ‘TwoSampleMR’ R package.^26-28^

This analysis had two aims: i) to further investigate the possibility of horizontal pleiotropy affecting the results of the eosinophil MR; and ii) to establish whether any other cell types besides eosinophils could affect the respiratory outcomes studied.

## Results

### Univariable MR analyses of eosinophils and respiratory outcomes

There were 151 SNPs available for the univariable MR analyses, which were used in analyses of three quantitative lung function traits (FEV_1_, FVC, and FEV_1_/FVC), and four respiratory disease phenotypes (moderate-to-severe asthma, AECOPD, ACO and respiratory infections). Details of the selection of SNP IVs is described in **Figure 1**.

**Figure 1.**
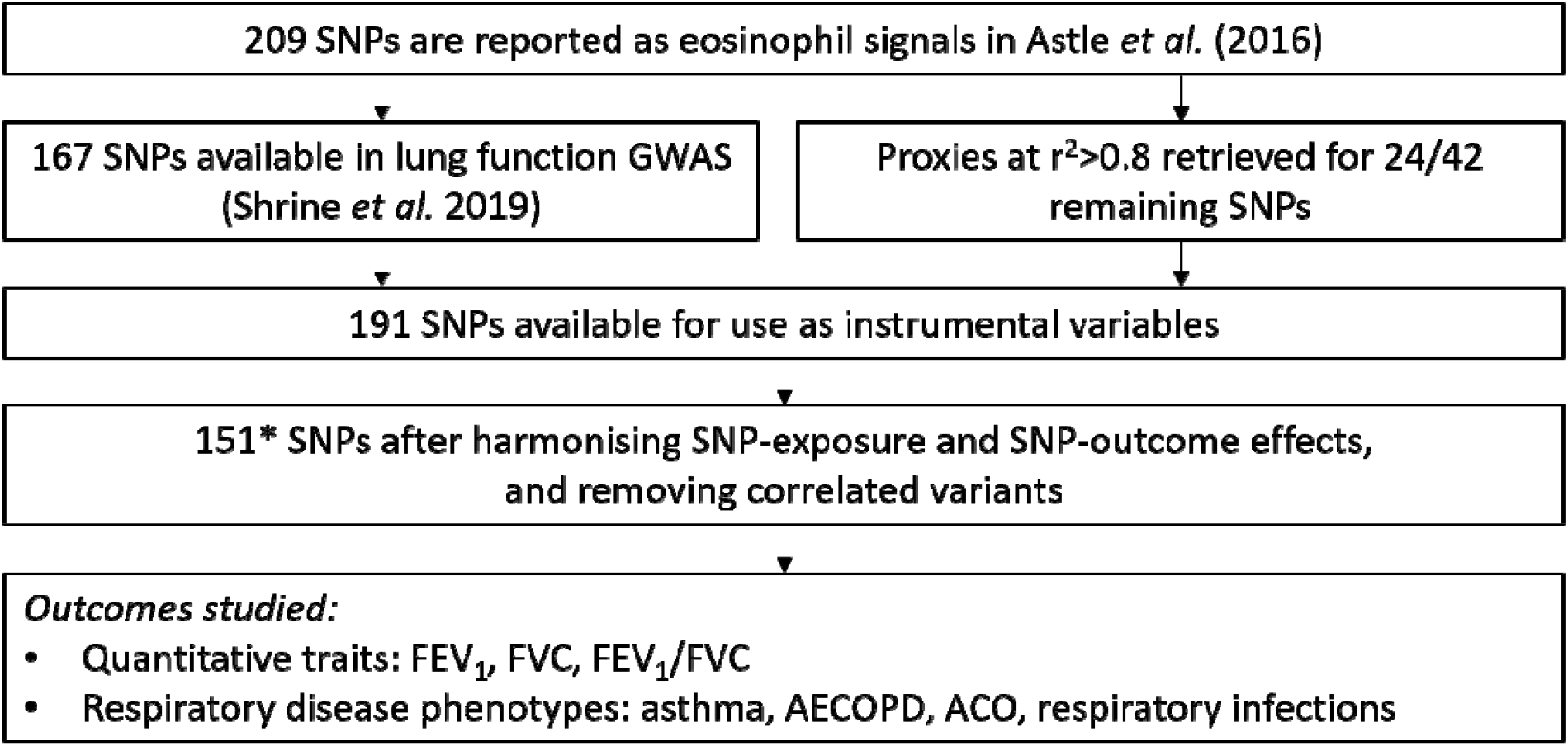
Selection of SNPs for univariable MR analyses of eosinophils and respiratory outcomes. Flowchart describing the analysis workflow for initial MR analyses of eosinophils. Of 209 SNPs associated with eosinophil count, 167 were available in lung function GWASs (missingness is due to some SpiroMeta studies not being imputed to the HRC panel) ^18^. LD proxies at r^2^ > 0.8 were retrieved for 24/42 missing variants. Of the resulting 191 SNPs, 188 were successfully harmonised between the SNP-eosinophil and SNP-lung function data sets, and 151* remained after LD clumping at an r^2^ threshold of 0.01. These 151 SNPs were used in analyses. *One SNP, rs9974367, was missing in the moderate-severe asthma GWAS. AECOPD=acute exacerbation of COPD; ACO=asthma-COPD overlap.

Results are presented in **Figure 2**. Amongst the quantitative lung function traits, there was strongest evidence for a causal effect of eosinophils on FEV_1_/FVC (SD change in FEV_1_/FVC per SD eosinophils, IVW estimate=−0.049 [95% CI: −0.079, −0.020]), with a smaller effect on FEV_1_ (IVW estimate=−0.028 [95% CI: −0.054, −0.003]). There was substantial heterogeneity of SNP-specific causal estimates for all three traits, as evidenced by the large values of Cochran’s Q statistic. However, heterogeneity was likely due to balanced pleiotropy, since weighted median estimates were consistent with the IVW estimates. Confidence intervals for the MR Egger estimates were broad, consistent with the low power of this method, but there was no evidence of unbalanced pleiotropy, as evidenced by the p-values for the intercept of the MR Egger intercept term. Scatterplots of SNP-outcome against SNP-exposure effects for the three quantitative outcomes are given in **Supplementary Figure 1**.

**Figure 2.**
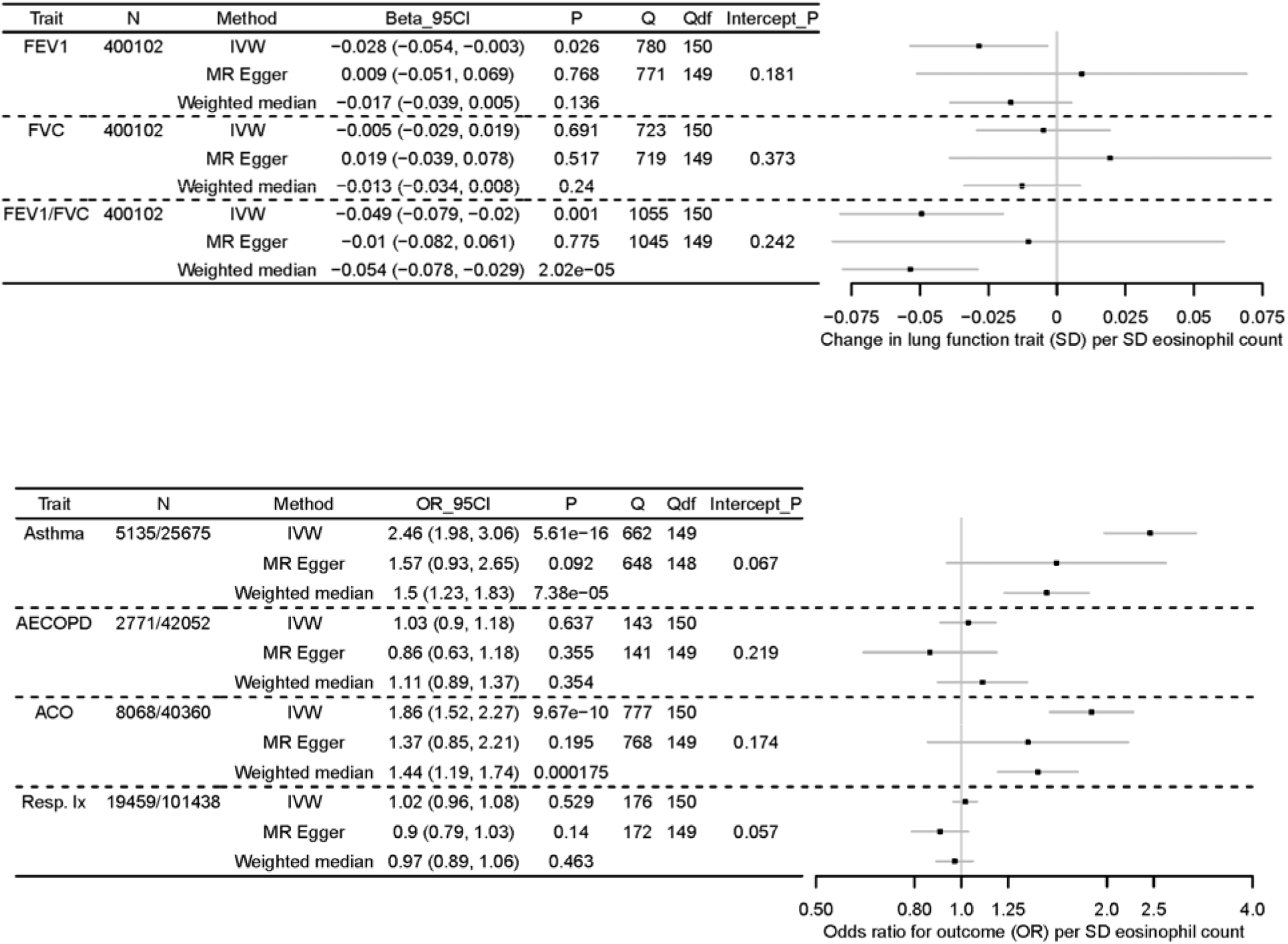
MR analyses of eosinophils (exposure) on three quantitative lung function traits (top) and four respiratory disease phenotypes (bottom), using 151 eosinophil-associated SNPs. Top: Results of MR analyses of eosinophil counts (exposure) on three quantitative lung function traits (outcome), FEV_1_, FVC, and FEV_1_/FVC. A forest plot of three estimates for each traits is shown (IVW, MR Egger, weighted median), along with the maximum sample size in the outcome GWAS (N), the effect size in SD change in outcome trait per SD eosinophil count, and 95% confidence interval, values for Cochran’s Q statistic (Q) and the associated degrees of freedom (Q_df), and the P-value for the MR Egger intercept (Intercept_P). Boxes of the forest plot represent effect sizes, whiskers are 95% confidence intervals. Bottom: Results of MR analyses of eosinophil counts (exposure) on four respiratory disease phenotypes (outcome), moderate-to-severe asthma, acute exacerbations of COPD (AECOPD), asthma-COPD overlap (ACO), and respiratory infection (Resp. Ix). A forest plot of three estimates for each traits is shown (IVW, MR Egger, weighted median), along with sample size in the outcome GWAS for cases and controls, respectively (N), the effect size as odds ratio (OR) per SD eosinophil count, and 95% confidence interval, values for Cochran’s Q statistic (Q) and the associated degrees of freedom (Q_df), and the P-value for the MR Egger intercept (Intercept_P). Boxes of the forest plot represent odds ratios, whiskers are 95% confidence intervals. NB only 150/151 of the eosinophil SNPs were available in the moderate-to-severe asthma GWAS.

Amongst the respiratory disease phenotypes (also **Figure 2**), there was strong evidence for a causal effect of eosinophils on asthma (OR per SD eosinophil count, IVW method=2.46 [95% CI: 1.98, 3.06]), and ACO (IVW OR=1.86 [95% CI: 1.52, 2.27]). There was substantial heterogeneity of SNP-specific causal estimates for these two traits, and the weighted median estimates were smaller in magnitude than the IVW estimates (weighted median OR: 1.50 [95% CI: 1.23, 1.83] for asthma, and 1.44 [95% CI: 1.19, 1.74] for ACO). Whilst the confidence intervals for the MR Egger estimates were still broad, estimates were generally closer to the weighted median estimates, suggesting that the IVW estimates were inflated due to the presence of horizontal pleiotropy, particularly for asthma. Inflation for the asthma estimate is also more likely due to larger overlap between the SNP-exposure and SNP-outcome datasets for this analysis (see **Extended Methods**). Scatterplots of SNP-outcome against SNP-exposure effects for all four outcomes are given in **Supplementary Figure** 2, which clearly demonstrate the heterogeneity in causal effect estimates. Conversely, there was no evidence of association of eosinophil counts with AECOPD or respiratory infections. For both outcomes, confidence intervals for all three MR estimates included the null, and all estimates were close to the null.

See **Supplementary Table 4** for full results for all models.

#### Sensitivity analysis to assess the effects of sample overlap for quantitative lung function traits

UK Biobank featured in all GWAS datasets used, albeit the blood cell count GWAS and asthma GWAS included only approximately one third of the UK Biobank genotype data.^14^ Since we had access to data from the subset of the quantitative lung function GWASs that did not include UK Biobank, we conducted sensitivity analyses to assess for the effect of sample overlap (see **Extended Methods**). Results were generally consistent with the main results (e.g. SD change in FEV_1_/FVC per SD eosinophil count, IVW estimate=−0.041 [95% CI: −0.072, −0.009]; SD change FEV_1_ per SD eosinophil count=−0.043 [95%CI −0.077,−0.010]) (**Supplementary Table 5**).

#### Sensitivity analysis to assess the effect on FEV_1_/FVC in individuals without asthma

The causal effect of eosinophils on FEV_1_/FVC was recalculated using data from UK Biobank, excluding individuals with an asthma diagnosis. The effect size was attenuated compared to the main results, and the 95% confidence interval overlapped the null: IVW −0.013 [95% CI: −0.041, 0.015] (see **Supplementary Table 6**).

### Multivariable MR analyses of blood cell counts and respiratory outcomes

To further explore causality between blood cell parameters and FEV_1_, FEV_1_/FVC, moderate-to-severe asthma and ACO, we carried out multivariable MR analyses, using eight cell types as exposures (basophils, eosinophils, neutrophils, monocytes, lymphocytes, platelets, red blood cells and reticulocytes).

The selection of SNP IVs for multivariable MR is described in the **Methods**, and **Supplementary Table 3**. Briefly, 1166 unique SNPs were associated with at least one of the eight cell types at p<8.31×10^−9^ in Astle *et al*., and were available in the outcome GWASs. After LD-clumping, 329 SNPs remained, and after harmonising SNP-exposure and SNP-outcome effects, 318 SNPs remained. Estimated variance explained (r^2^) and instrument strength (F-statistics) by cell-type are also given in **Supplementary Table 3**. We present these estimates as a guide of relative strength of sets of IVs, but do not intend for them to be interpreted as measures of absolute strength, given the likelihood of Winner’s curse bias.^20^

Multivariable MR results for FEV_1_ and FEV_1_/FVC are presented in **Figure 3**. Even after conditioning on the effects of the SNPs on other cell types, eosinophils reduced lung function as measured by FEV_1_/FVC (multivariable estimate, SD change in FEV_1_/FVC per SD eosinophils adjusted for other cell types: −0.065 [95% CI: −0.104, −0.026]). The eosinophil point estimate for FEV_1_ was consistent with the univariable estimate (**Figure 2**), but results for eosinophils (−0.032 [95%CI: −0.068, 0.005]) and other cell types were consistent with the null. When asthma cases were excluded from SNP-FEV_1_/FVC results, the eosinophil estimate attenuated, and confidence intervals overlapped the null (−0.028 [95%CI: −0.069, 0.013]) consistent with the causal effect of eosinophils on lung function being of greater magnitude in people with a history of asthma (**Supplementary Figure 3**).

**Figure 3.**
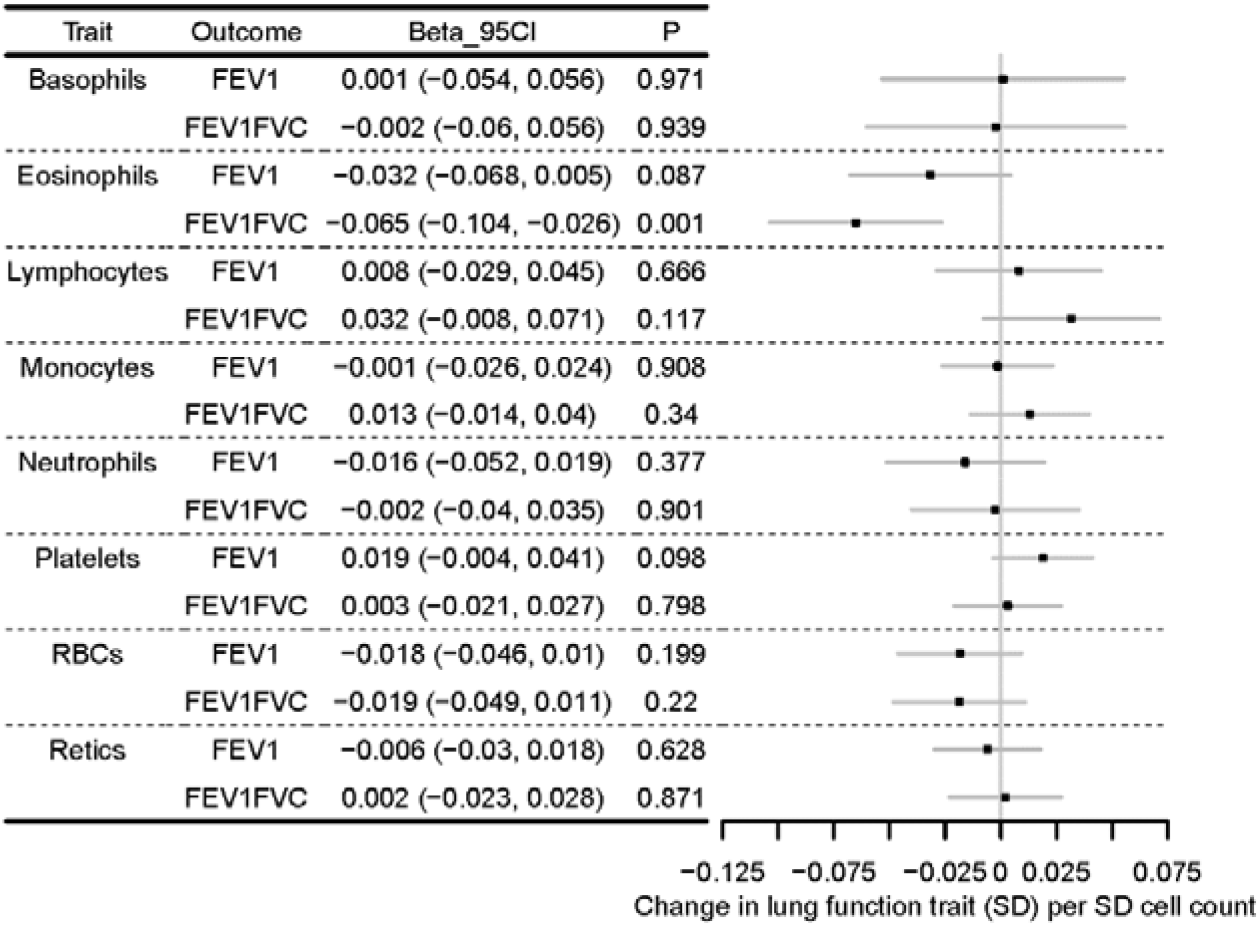
Multivariable MR analyses of eight cell types and FEV_1_ and FEV1/FVC. Forest plot showing multivariable MR estimating the causal effect of multiple cell types on quantitative lung function outcomes, after conditioning on the effects of the SNPs on other cell types. Models were run for each of forced expiratory volume in 1 second (FEV_1_) and the ratio of FEV_1_ to forced vital capacity (FVC) separately, but effect sizes are shown next to one another for comparison. Effect sizes (beta, 95% confidence interval, 95CI) are in SD change in lung function outcome per SD cell count (adjusted for the effects of other cell types). Points of the forest plot represent effect size estimate; whiskers are 95% confidence intervals.

Results of the multivariable MR analysis for the two respiratory disease outcomes (ACO and asthma) are presented in **Figure 4**. There was an association of eosinophil count with both ACO (OR 1.95 [95% CI: 1.57, 2.42]) and asthma [OR 2.90 [95% CI: 2.31, 3.65]], after adjusting for the effects of the SNPs on other cell types. Confidence intervals for other cell type estimates were consistent with the null effect of 1, with the exception of neutrophils for ACO.

**Figure 4.**
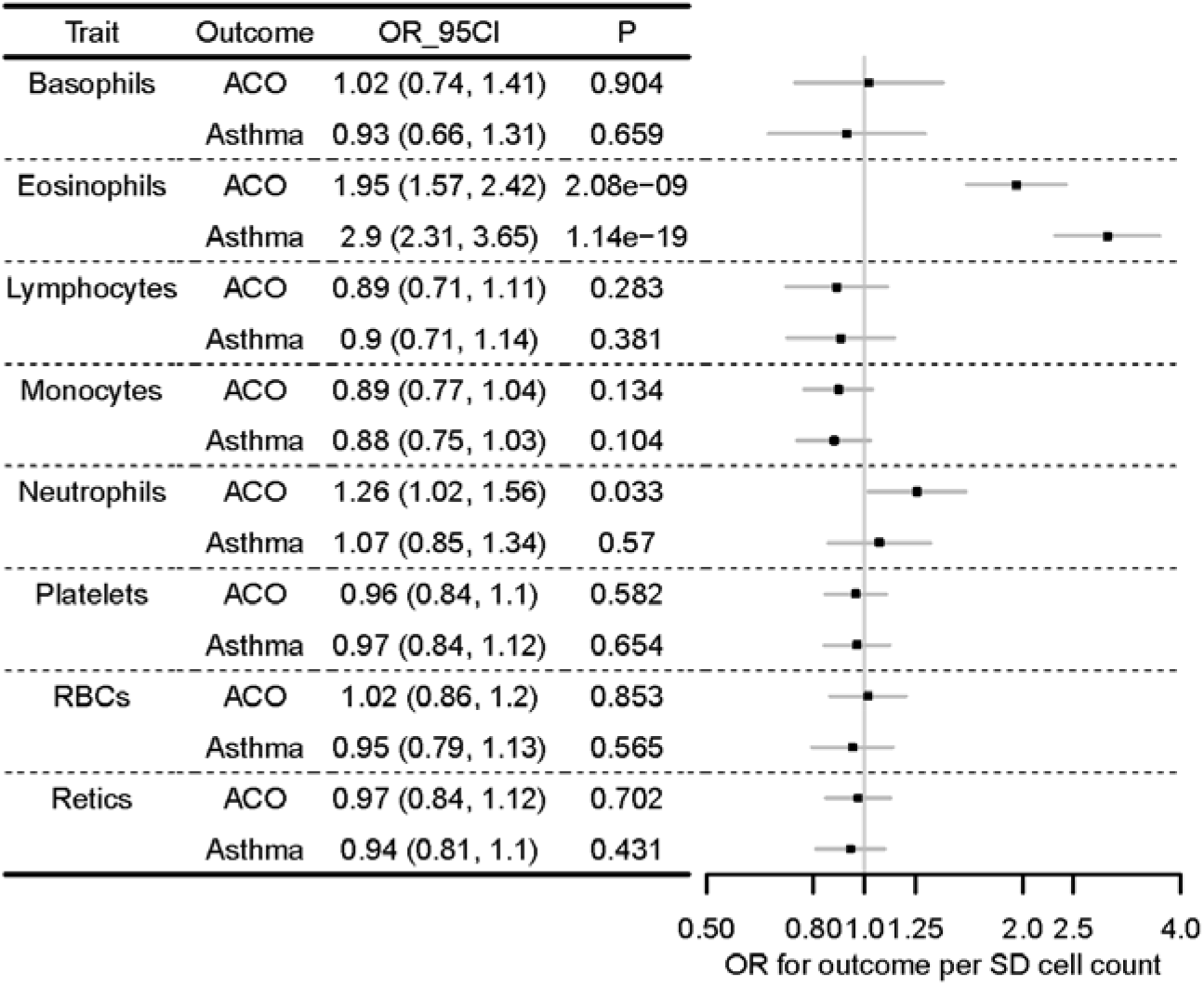
Multivariable MR analyses of eight cell types and two respiratory disease outcomes, ACO and asthma. Forest plot showing multivariable MR estimating the causal effect of multiple cell types on respiratory disease outcomes, after conditioning on the effects of the SNPs on other cell types. Models were run for each of ACO and asthma separately, but effect sizes are shown next to one another for comparison. Odds ratios (OR, 95% confidence interval, 95CI) are per SD cell count (adjusted for the effects of other cell types). Points of the forest plot represent odds ratios; whiskers are 95% confidence intervals.

## Discussion

Using Mendelian randomization, we found evidence for a causal effect of eosinophils in determining FEV_1_/FVC, FEV_1_, ACO and asthma. For these four outcomes, we additionally performed multivariable MR to investigate the causal role of a broader range of eight blood cell types, and found that amongst these cell types eosinophils had the strongest evidence of an effect on the respiratory outcomes studied.

To our knowledge, this is the largest MR study of eosinophils in relation to lung function, and the first to investigate the relationship between eosinophils and AECOPD, ACO, and respiratory infections. Whilst terminology of ACO has changed over time, the recognition that asthma and COPD coexist in some patients has not changed,^29^ and this is what our analysis aimed to capture.

A previous two-sample MR analysis of eosinophils in relation to asthma was undertaken by the authors of the cell count GWAS used to derive IVs in the current study; these authors used asthma GWAS data from the GABRIEL study.^14^ Our results support their conclusion that eosinophils are causal determinants of asthma. We are aware of one other MR study of eosinophil counts in relation to asthma, COPD, FEV_1_ and FEV_1_/FVC, conducted entirely within the LifeLines cohort (N=13,301, 5 SNPs used as IVs).^16^ In that study, confidence intervals for causal estimates of eosinophils were consistent with the null, albeit point estimates were consistent with a harmful effect for FEV_1_/FVC, asthma and COPD. Here, we used the largest eosinophil count GWAS to date (N=172,275)^14^ to derive IVs, and found evidence for causality of eosinophils in reducing FEV_1_/FVC, the lung function trait in which impairment is the key feature of COPD diagnosis, and FEV_1_, the trait used to grade airflow limitation in COPD. We did not find strong evidence of a causal effect of eosinophils on FVC, despite observing negative point estimates. This is of interest because it is a reduction in FEV_1_ (and therefore in FEV_1_/FVC) that is a hallmark of obstructive lung diseases, with effects on FVC being generally less prominent. Our sensitivity analyses highlighted a larger estimated causal effect of eosinophils on FEV_1_/FVC among individuals with a history of asthma, with an attenuation of effect size estimates seen when excluding this group. Together with the effect shown for ACO, our findings highlight the importance of eosinophils on lung function and airflow obstruction in people with a history of asthma.

Amongst the outcome traits showing evidence for causation by eosinophils, we observed substantial heterogeneity in causal estimates for individual SNPs, i.e. different causal effects on the outcome phenotypes may arise from different genetic variants influencing eosinophils. Accordingly, we compared MR methods that rely on differing assumptions for validity (**Box 1**). When comparing the IVW, MR-Egger and weighted median results, effect estimates for eosinophils on quantitative lung function traits were broadly consistent, but point estimates for ACO and asthma attenuated substantially when using the MR-Egger and weighted median approaches (albeit estimates were still consistent with a harmful effect of raised eosinophils). This suggests there were SNPs with pleiotropic associations, e.g. SNPs associated with asthma and ACO via pathways other than eosinophils, which is a known challenge in MR studies (see also **Box 1**).

Since many of the SNPs used as IVs for eosinophil count are also associated with other cell counts,^14^ we performed multivariable MR to estimate the influence of multiple cell types simultaneously, after conditioning on the effects of the SNPs on other cell types. Whilst such an analysis has previously been undertaken for asthma,^14^ we also performed multivariable MR analyses for FEV_1_, FEV_1_ /FVC, asthma and ACO. Whilst we did not find substantial evidence for a harmful effect of neutrophils on asthma, nor a protective effect of monocytes and lymphocytes, as reported by Astle *et al*.,_14_ the effect directions in our multivariable MR analysis were consistent with those of this previous study for neutrophils, monocytes and lymphocytes. In multivariable MR analyses, we observed a larger effect of eosinophils on asthma than reported previously: this could be because our SNP-outcome dataset was of moderate-to-severe asthma, and around half of the cases and the majority of controls were also included in the exposure GWAS. Nevertheless, the univariable MR estimates from MR-Egger regression and weighted median estimation were consistent with the previous estimate reported for asthma in multivariable analysis by Astle *et al*.^14^

In comparison with the harmful effect of eosinophils on FEV_1_/FVC, we did not observe a harmful effect of neutrophils on FEV_1_ /FVC, despite the known relationship between neutrophilia and COPD.^30^ This could be due to lower power of the neutrophil IVs compared to the eosinophil IVs (**Supplementary Table 3**), but it is also possible that neutrophilic inflammation is more of a consequence than a cause of reduced FEV_1_ /FVC, for example in response to infection,^31^ and therefore may represent a marker of disease progression. There was very weak evidence for a harmful effect of neutrophils on ACO. Whilst we did not find strong evidence for causality of eosinophils on AECOPD and respiratory infections, point estimates were consistent with a harmful effect on AECOPD, and may have been limited by power. The effects of anti-IL5 drugs that have been attributed to the reduction of eosinophils have been noted to be smaller in acute exacerbations of COPD compared to asthma.^3,32^

Key strengths of the study are that we carried out two-sample analyses with differing sensitivities to underlying assumptions (including MR-Egger and weighted median analysis). We used the largest GWAS dataset available for eosinophil count together with multiple large GWASs of lung function and respiratory disease. In addition to the advantages of valid MR analyses in mitigating against some key limitations of observational epidemiology (reverse causality and confounding), using genetic proxies for eosinophils may provide a better estimate for long-term eosinophilia, which may be useful, since one-off assays of cell counts may show a degree of lability.^33^ Another strength of our work is that we undertook multivariable MR analyses to investigate causality between multiple cell types and the outcomes studied, whilst controlling for the effects of IVs that may have had pleiotropic effects via other cell types.

We acknowledge several limitations. We did not have post-bronchodilator measures of spirometry available. However, our criterion for defining asthma-COPD overlap required a % predicted FEV1 of less than 80% (consistent with a spirometric definition of GOLD Stage 2-4 COPD); using the same pre-bronchodilator spirometry definition of COPD we have previously shown a positive predictive value of 98% for diagnosis of postbronchodilation-defined COPD.^34^ Sample overlap between the SNP-eosinophil and SNP-outcome datasets (all datasets included participants from UK Biobank) could bias estimates towards the observational eosinophil-outcome association.^20^ As a sensitivity analysis, we repeated the univariable lung function MR analysis of eosinophils using SNP-lung function results from the SpiroMeta consortium (i.e. omitting UK Biobank), and found a consistent IVW estimate. Nevertheless, we acknowledge that our analyses of other outcomes (particularly the asthma analysis) could be vulnerable to a degree of non-conservative bias.^19,20^ The results of our MR analyses also use genome-wide results adjusted for covariates, and therefore may be susceptible to collider bias.^19^ For the binary phenotypes of asthma, AECOPD and respiratory infections, there may also have been bias due to misclassification in electronic healthcare records, which we would expect to be non-differential, and therefore towards the null. There is also potential bias in the causal estimates for binary outcomes due to non-collapsibility of the odds ratio.^21^ Our analyses do not consider the possibility of non-linear effects between eosinophil counts and lung function, which have been observed: the results presented are the average difference in lung function if eosinophils were increased by the same amount for every individual in the population. Finally, we acknowledge that the multivariable analyses may still be vulnerable to residual pleiotropy via pathways other than the eight cell types studied. However, we argue that the sensitivity analyses undertaken together support a causal role for eosinophils in the traits studied. The significant heterogeneity in SNP-specific causal estimates may arise from pleiotropy. A specific genetic variant influencing eosinophils may have a greater or lesser effect on these respiratory phenotypes than the direct effect of eosinophils, possibly in the opposing direction.

Our work provides insight into the role of eosinophils in causing respiratory conditions characterised by airflow obstruction. At present, treatment with the anti-IL5 agents mepolizumab and benralizumab in asthma is initiated according to levels of eosinophil counts,^9^ yet it is possible that a more proximal factor, potentially such as IL5 itself, may be an even better predictor of drug response. Future work could seek therefore to identify whether particular pathways upstream of eosinophil counts might help design better methods for deciding upon treatment initiation. To conclude, we found new evidence for a causal effect of eosinophils on ACO, FEV_1_, and FEV_1_/FVC, as well as reproducing a previous association with asthma. Taken together, our findings support eosinophils being important causal determinants of airflow obstruction, and suggest that the strongest causal effects of eosinophils are in people with a history of reversible airflow obstruction, including the patient group with features of both asthma and COPD.

## Data Availability

This manuscript uses summary-level GWAS data. Lung function GWAS summary statistics as well as the blood cell GWAS summary statistics are available from the EBI GWAS catalog (https://www.ebi.ac.uk/gwas/). Other GWAS summary statistics are available from the authors on request.

## URLs

EBI GWAS catalog: www.ebi.ac.uk/gwas

rAggr: http://raggr.usc.edu/

‘TwoSampleMR’ R package documentation: https://mrcieu.github.io/TwoSampleMR/

## Author Contributions

NS, ALG, ATW, CJ, NR, the SpiroMeta consortium, IPH, LVW and MDT were involved in data collection, by generating GWAS data of the respiratory traits used in these analyses. NAS, FD, IPH and MDT were involved in data interpretation. IPH and MDT were involved in study design. ALG was involved in all aspects of the work, performed the analysis and wrote the first draft of the paper, including all figures and tables. All authors critically reviewed the manuscript and contributed to data interpretation and draft revision.

## Ethics committee approval

This research used published summary data from published genome-wide association studies, plus data from UK Biobank. This particular research was conducted using the UK Biobank Resource under Application Number 648. UK Biobank has ethical approval from the UK National Health Service (NHS) National Research Ethics Service (Ref 11/NW/0382).

## Acknowledgements

We are grateful to Dr William J Astle for his advice. This research used the ALICE and SPECTRE High Performance Computing Facilities at the University of Leicester. This analysis used summary data available via the BREATHE Digital Innovation Hub hosted at SAIL databank (https://saildatabank.com/).

